# Arachnoiditis: leveraging crowdsourcing and AI in a cross-sectional study of 1,105 cases to improve identification, understanding, and treatment

**DOI:** 10.1101/2025.11.18.25340508

**Authors:** Lori Verton, Noga Minsky, Eran Dotan, Roni Sharon, Megan Black, Patricia Gomes, Deepmala Rana Bhat, Shristi Sharma, Ishna Singh, Linda Bavisotto

**Author notes:** **Correspondence:** Linda Bavisotto.

## Abstract

**Background:** Arachnoiditis, a painful and potentially disabling neurological condition, results from persistent inflammation of the spinal cord pia-arachnoid membranes following injury. While considered rare, the condition is underdiagnosed. Research on symptomatology, diagnosis, and treatments is scarce, hindering clinical management. Artificial intelligence (AI) offers promising opportunities for rare diseases, enabling large-scale pattern identification. This study used traditional research methods coupled with AI technology to characterize patient-reported clinical presentation, comorbidities, aggravating factors, and treatments for arachnoiditis.

**Methods:** This retrospective cross-sectional study utilized data from StuffThatWorks (STW), an online crowdsourcing platform for people with chronic diseases. Multiple choice and free text responses were assessed both quantitatively and qualitatively. Novel AI/machine learning algorithms were used to further analyze the data, including the STW cross-condition score (higher scores more indicative of arachnoiditis) and the STW treatment efficacy model generating effectiveness and detriment estimates, with binomial proportion 90% confidence intervals.

**Results:** Of 1250 international participants, 1105 reporting a physician-confirmed diagnosis were included. Participants were predominantly USA-based (71.4%), female (75.9%) and ≥46 years old (73.1%). Of 712 symptoms grouped into eight categories, eighteen were more indicative of arachnoiditis (by cross-condition score). The most frequent symptoms were lower back pain (43.5%), leg pain (41.6%) and back pain (39.1%). Prolonged sitting (62.5%) and prolonged standing (58.3%) were the most common aggravating factors. Comorbidities were led by degenerative disc disease (32.3%), spinal stenosis (25.3%) and fibromyalgia (25.0%). The most frequently used treatments were gabapentin (37.9%), physiotherapy (30.1%) and pregabalin (26.5%). Treatments with the highest patient-rated effectiveness (by STW model, 90% CI) were low-dose naltrexone (28.1%, CI 20.0-37.0), ketamine infusion (24.8%, CI 16.9-33.4) and fentanyl (21.1%, CI 14.7-28.1). Epidural corticosteroid injections showed the highest detriment (38.5%, CI 28.0-45.9).

**Conclusion:** As the largest observational study of arachnoiditis to date, made possible with novel methodological approaches, this work offers new insights with potential to improve diagnosis and management.

## Background

Arachnoiditis is a painful, potentially debilitating neurological condition resulting from persistent inflammation of the spinal cord pia-arachnoid membranes (leptomeninges) and the subarachnoid space between them [1–3]. A cascade of neuroinflammation in response to injury—a normally healthy mechanism, but persistent or excessive in arachnoiditis—results in early spinal nerve swelling and leads to thickening and clumping of spinal nerve roots [1,3,4]. The process can be focal or diffuse, and over time becomes compounded by the development of permanent scarring and adhesions, entrapping nerves and disrupting the normal flow of cerebrospinal fluid (CSF) in the subarachnoid space. Fibrosis and CSF flow disturbances can lead to spinal cord tethering, syrinx formation, myelomalacia, spinal cord atrophy, and myelopathic symptoms [1,2,5–9].

Arachnoiditis manifests with a wide spectrum of clinical presentations, and the course of the condition is highly variable [10]. Advanced stage adhesive arachnoiditis is commonly characterized by intractable neuropathic pain, significant mobility issues and myriad neurologic impairments [1,9,11]. Complications can extend to the genitourinary, gastrointestinal and autonomic nervous systems [12,13]. Together, these disease expressions can lead to significant morbidity, disability and shortened lifespan [3,13,14]. Considered a rare condition, but often misdiagnosed and overlooked, much remains unknown about the epidemiology and natural history of arachnoiditis, or the efficacy of interventions [1,15].

Clinical symptoms of arachnoiditis can appear acutely within hours or days of an inciting event, or develop gradually over months to years, or even decades, often leading to delayed diagnosis or misdiagnosis [1,9,11,16]. The condition may be triggered by a variety of factors, primarily involving physical damage or chemical injury to the spinal cord, or by infection of the spinal meninges. Injury may occur because of trauma to the spinal cord, subarachnoid hemorrhage or other introduction of blood into the CSF, edema, or post-surgical or post-procedural inflammation [1,5,17,18]. Exposure of the meninges and CSF to a variety of chemicals has also been documented to cause arachnoiditis, with offending agents including intrathecal chemotherapy, corticosteroids, oil-based radiographic contrast agents, detergent solvents, contaminants, and preservatives such as polyethylene glycol, to name just a few [1,19]. Infectious meningitis can be caused by fungal, bacterial, viral, or parasitic pathogens [1,20]. Chronic meningitis, or aseptic arachnoiditis, are terms sometimes used to refer to inflammation without evidence for a causal infectious pathogen. The condition can also be triggered by autoimmunity, for example as an adverse immune event due to immune checkpoint inhibitor therapy of various malignancies [21]. In addition, idiopathic cases are not uncommon [1,22].

Whereas arachnoiditis arising as a late complication of meningeal infection may favor cervical and thoracic spine distribution [1], where the disease tends to be more progressive [2], the current predominance of lumbar arachnoiditis appears to be related to an increasing frequency of invasive lumbar spinal interventions, including various surgical techniques as well as epidural anesthesia and injections. Iodinated oil-based myelographic contrast imaging agents, such as iophendylate, administered via lumbar puncture into the subarachnoid space, have been associated with the development of numerous cases of iatrogenic arachnoiditis [1,4,14,19], and these agents were subsequently removed from the market. Fungal meningitis and arachnoiditis were documented in a number of clustered cases arising from contamination of methylprednisolone in epidural injections in 2012 [23–25] and of spinal anesthesia in 2023 [26]. An updated American practice advisory was issued in 2017 on the prevention, diagnosis and management of infectious complications associated with neuraxial techniques [27].

It is important to emphasize that most individuals with the aforementioned exposures do not develop arachnoiditis. Some individuals may have underlying risk factors that predispose them. There appears to be an association with pre-existing anatomic spinal abnormalities as well as age-related degeneration, particularly in lumbar arachnoiditis [3,7,14,28,29]. These conditions in turn also lead to increased invasive spinal interventions, with multiple spinal interventions (of any kind) or traumatic procedures being more closely associated with the development of arachnoiditis [1,4,30]. There may be an increased risk in persons with auto-immune or systemic inflammatory conditions, such as autoimmune vasculitis, ankylosing spondylitis, mixed connective tissue disease or rheumatoid arthritis [1,31–33]. Additionally, a potential association with genetic connective tissue disorders, such as Ehlers-Danlos Syndrome (EDS), has been described in a limited number of studies [34,35]. Rare cases of familial arachnoiditis with syringomyelia have been documented [36], suggesting the possibility of an unidentified genetic susceptibility component in these individuals. A potential association between arachnoiditis and Tarlov (perineural) cysts has also been reported [34,37,38]. Various authors suggest that, in many cases, arachnoiditis is likely to result from a combination of risk factors or inciting events [4,14,34,39].

Diagnosis of arachnoiditis is based on individual history, clinical findings and MRI features. MRI is the preferred imaging modality, although CT myelography may be used when MRI is contraindicated [29]. Over the past 50 years, several staging classification systems for arachnoiditis have been proposed and modified based on symptoms, histopathology, morphology and patterns of radiographic progression [14,40,41]. Expanding upon the pathognomonic radiological patterns first established in the 1980s [40,42], recent diagnostic studies of advanced stage adhesive arachnoiditis show that MRI findings may include abnormal clumping and irregular distribution of spinal nerve roots, abnormal nerve root contours, nerve root enhancement, synechia, loculated and typically dorsal arachnoid cysts, as well as signs of CSF flow disturbance [10,28,29,43]. While radiological staging classifications have focused primarily on lumbar arachnoiditis, disease occurring in other spinal areas and the brain may be even more difficult to characterize on MRI [44–46].

Several limitations complicate the diagnosis of arachnoiditis in clinical practice. Imaging findings are highly variable [29], and correlate poorly with symptom severity [43,47]. In the cervicothoracic spine, neurosurgical observations show that MRI often incompletely visualizes the extent of chronic arachnoiditis, especially focal arachnoiditis [44,45]. In the lumbar spine, endoscopic surgery suggests that, in some cases, MRI may fail to identify the condition at all [37,48,49], leading some authors to propose that mild arachnoiditis may be overlooked in patients with unexplained lumbar radiculopathy symptoms but normal MRI studies [50]. Adding to these diagnostic challenges, acute iatrogenic lumbar arachnoiditis may initially present with minimal MRI findings – early imaging may show only nerve root enhancement before other characteristic features develop, or may appear entirely normal [51–55].

To address these limitations, several diagnostic methods have recently been described. In the cervicothoracic spine, specialized MRI sequences and morphometric analyses for improving visualization or inferring diagnosis have been employed [44,56–59]. In the lumbar spine, proposed approaches include prone MRI to detect failure of the nerve roots to migrate ventrally with gravity due to nerve root tethering [60], and morphometric analysis to assess for mildly enlarged thecal sac dimensions [22,48,61,62]. MR neurography of the lumbosacral plexus has also detected arachnoiditis not otherwise apparent on standard MRI [63]. In the absence of a definitive diagnostic test and reflecting a lack of clear consensus in the field, Epstein and Agulnick [47] have suggested that a characteristic constellation of symptoms alone should suffice to make the clinical diagnosis of lumbar arachnoiditis, without the requirement for radiographic confirmation.

Early and accurate diagnosis is important, as some therapeutic options for arachnoiditis may be more effective when given in the initial stages of neuroinflammation. When the diagnosis is not definitive, however, early intervention may be delayed to avoid possible complications. In those with recent onset, early stage arachnoiditis, a variety of anti-inflammatory, fibrinolytic, and immune-suppressive therapies have been empirically offered in an attempt to arrest or slow the process of fibrosis and neuropathic progression [1,12,53,64,65]. According to several recent reports, high-dose corticosteroids administered in the earliest stages of acute arachnoiditis, often in iatrogenic cases, have led to recovery in the short term [52,53,64,66,67]. However, the evidence is mixed, not all patients respond or respond completely, and some patients recover spontaneously [1,52]. Most reports are missing sufficient details. Further studies with standardized treatments and long-term outcome assessments are needed as a critical research priority.

In patients experiencing progressive neurological deterioration, surgical intervention such as detethering procedures, adhesiolysis, cyst fenestration, and decompression of intradural arachnoid cyst or syrinx with or without shunting may be indicated [1,2,5,44,62,68,69], however these procedures carry inherent risks of potential further compromise in neurological function as well as development of post-operative inflammation and scarring, which may lead to arachnoiditis recurrence. The primary surgical objectives are to restore or improve CSF flow at sites of obstruction and to alleviate spinal cord compression.

The majority of contemporary literature on surgical management report on cases of thoracic and cervical arachnoiditis, although some have included lumbosacral cases [5]. Favorable surgical outcomes have been reported, particularly early in disease prior to significant spinal cord damage [2], although follow-up is often short and information incomplete. There is relatively consistent evidence indicating that patients with focal spinal involvement of arachnoiditis (limited to several vertebral segments) have better surgical and neurologic outcomes post-operatively than those with extensive/diffuse spinal involvement [2,44]. Minimally invasive neurosurgical approaches such as percutaneous intrathecal endoscopy continue to evolve [37,48,49,62,70,71] and are sometimes combined with limited laminectomy procedures [2,69] to allow direct visual assessment and safe removal of adhesions in spinal segments adjacent to the laminectomy margins, extending the range of the surgeon’s reach beyond what would be possible with conventional laminectomy.

For patients who are not surgical candidates, management focuses on symptom-directed supportive care and comprehensive chronic pain management [72,73]. Current pharmacological and non-surgical interventions have variable results, typically providing modest symptom relief at best, often with limited long-term benefit [1,3,11,12].

Substantial hurdles have impeded studies regarding the diagnosis, natural history and treatments for arachnoiditis. One challenge is the lack of standardized terminology for arachnoiditis across its varying degrees of severity, with Palackdharry et al. identifying up to 63 different terms used across specialties [1]. Determining clinically significant pathology also remains unclear [74,75]. In addition, because arachnoiditis spans multiple specialties without fitting neatly within one, research is fragmented [74]. Consequently, limited literature exists regarding pathophysiology, symptomatology and accurate diagnosis, rendering correct identification of the condition difficult. Inaccurate diagnosis may be common, as reflected by poor inter-rater reliability (kappa score 0.05) in a 2021 study where five specialized radiologists reviewed MRI images from 96 patients with hallmark criteria of the disease [28]. The inconsistent lag times between insult and clinical presentation may further contribute to delayed and missed diagnosis [10,61]. Chronicity of symptoms and the presence of comorbidities may also complicate the clinical picture.

In recent years, the advancement of bio-registries, online patient advocacy groups, digital health platforms, and disease registries, along with big data analytics, have begun to overcome financial and logistical hurdles to understanding and treating rare diseases. In addition, there has been increasing interest in arachnoiditis, with the number of publications growing [1,76]. Following a collaborative research network model in which patient-led initiatives have successfully accelerated research into rare diseases [77–78], the Arachnoiditis and Chronic Meningitis Collaborative Research Network (ACMCRN) [79] advocated for the establishment of an arachnoiditis research community on the digital health platform StuffThatWorks (STW) [80]. The platform is structured to enable community-driven research and insights. This study represents a retrospective analysis of the data accumulated through this initiative in order to help advance understanding of the condition. It characterizes the symptoms, aggravating factors, comorbidities and treatments among a large cohort of individuals living with arachnoiditis. This work serves as a prelude to the creation of an international arachnoiditis patient registry and the development of further studies.

## Methods

### Study design and data collection

This study is a retrospective cross-sectional analysis of observational patient-reported data collected by STW [80]. STW is a free commercial crowdsourcing digital health platform with approximately 2.9 million members across 1,261 medical conditions at the time of this writing. It provides an online venue for people living with specific medical conditions to voluntarily share structured information about their symptoms, treatments and outcomes within condition-specific groups, termed “communities”. This information is de-identified and aggregated into community insights that form an accessible organized knowledge database, enabling community members to self-educate. Natural language processing and machine learning algorithms aggregate and analyze entries to generate insights about age of onset, symptoms, aggravating factors and treatments. After several hundred responses are collected, the algorithm ranks treatments according to effectiveness; with larger numbers of responses, the model estimates the probability of treatment effectiveness based on user assessments.

Participation in the STW online survey platform is entirely voluntary, confidential and anonymous, and participants retain the right to opt out of the platform and data sharing at any time. Each of the STW chronic disease communities utilizes a common English-language, structured medical questionnaire. This includes demographic items as well as aspects of medical history, such as symptoms, diagnosis, treatments, aggravating factors, clinical indicators, and comorbidities, along with assessments of quality of life and daily functioning. With the exception of several mandatory questions, including current age and sex at birth, participants are free to disclose only information they want to share.

For the purposes of the current retrospective study, survey data were accessed in January 2023 and provided to the authors in aggregate format, with age categories, rather than precise age. Compiled data comprised participant responses to a questionnaire, including current age, sex at birth, age at symptom onset, symptoms, aggravating factors, comorbid conditions, and all previously attempted arachnoiditis treatments (including medicines, supplements, procedures, protocols, dietary changes, alternative or complementary therapies, classes). Supplementary Material 1 lists the specific STW survey questions used in the data analysis for this study. Many survey questions used open-text fields so as not to limit responses to a predetermined list. This exploratory approach necessitated that compiled results include both qualitative and quantitative data.

### Participants

Participants were users of the STW platform who voluntarily joined the condition-specific arachnoiditis community through multiple pathways, including organic discovery as well as promotion by ACMCRN [79]. ACMCRN, a patient advocacy and research organization, raised awareness of the arachnoiditis research community on the STW platform by drawing on its existing networks through social media and word-of-mouth. ACMCRN had no financial arrangement with STW and offered no incentives to participate on the STW platform. To be included in the study analysis, participants had to indicate that their arachnoiditis was diagnosed by a physician.

### Data analysis

Responses to open-ended questions were coded into structured entities using STW’s proprietary “normalization tool” as a component of natural language processing. For example, for a question regarding main symptoms, the answer “leg weakness and tingling” would be normalized into two different “symptom” entities: “leg weakness” and “leg tingling”. Numbers for each response were given out of the total number of valid responses (n), after exclusion of patients who did not answer the question and responses that were not normalized successfully by the normalization tool.

Quantitative data are reported as frequencies and percentages. Quantitative results also include values derived from an innovative algorithm unique to STW, the proprietary “STW cross-condition score”. This score uses AI-based models to measure how specific an entity or response is to one disease community when compared to all other chronic disease communities on the platform. The scale ranges from 1-10, with a score of 10 for an entity (such as a symptom, aggravating factor or comorbidity) which is very commonly reported in one community (e.g., arachnoiditis) but is rarely reported in others. Thus, higher scores indicate higher specificity for the condition of interest. A cross-condition score ≥ 6 is highly indicative of a given condition; a score between 3-5 is moderately indicative for the condition; and a score < 3 is a weak indicator for the condition. Novel research approaches are increasingly recognized as important for advancing understanding of rare diseases [81]. Given that results from other STW analytical models have demonstrated consistency with traditional research findings [82–83], the authors included the cross-condition score data in addition to standard research results.

To evaluate treatment effectiveness, alongside the primary data, a proprietary STW treatment efficacy model was used [82–84]. Treatment effectiveness was expressed as an estimated ratio of how many respondents rated a treatment as effective, out of the total number of reports of this treatment (i.e. number of effective reports/numbers of tried reports), while accounting for the level of certainty in the estimate based on the overall number of reports for each treatment. A hierarchical Bayesian calculation generates a binomial proportion confidence interval around the estimated proportion. To prevent overestimation of the effectiveness of rarely used treatments, the model accounts for the number of reports on each treatment, such that treatments with many reports are given greater weighting than those with few reports. The model does not distinguish between treatments which were taken for specific symptoms but rather assumes that all treatments are targeting the condition as a whole. The output is expressed as a percentage together with a 90% confidence interval.

Where a large volume of findings was captured from open-ended questions, results are presented according to a combination of both quantitative and qualitative methods. Qualitative analysis utilized content analysis methodology and categorization [85] of like responses for symptoms and aggravating factors, where ≥ 1% of the total study population entered the same response. This analysis enabled a detailed exploration of the full breadth of responses. In the absence of substantial prior research, qualitative methods are critical for generating insight, and, in the context of rare diseases, mixed method approaches are valued for incorporating lived experiences as meaningful input [86–87]. Although three authors (MB, LB, IS) did the content analysis and categorization, all authors agreed upon the final categorization of qualitative data.

The STW proprietary tools used in this study were applied within the platform environment. Subsequent to the data extraction performed by STW, the aggregated outputs generated by these models were shared with the authors, who have further analyzed and interpreted them in this report. No modifications to the underlying algorithms were made for the purposes of this study.

At the conclusion of the project, ACMCRN offered a summary of aggregate study findings and insights to the arachnoiditis online support groups community in a series of publicized online webinars. The sharing back of results to the participant community reflects recognized ethical principles of reciprocity and respect for participants’ contribution to the research.

### Ethics and data protection

Participation in the STW platform is voluntary, and the platform is designed to be transparent to its users, including in its use of artificial intelligence. Individuals join to share experiences related to specific health conditions and benefit from access to insights generated from their aggregated community data. At registration, before sharing any data, users agree to the platform’s terms of use and privacy policy, which inform participants that user-contributed information is handled in de-identified form and may be used for research purposes, including secondary analysis by external researchers. The policies specify that any data shared externally will be provided in aggregated form only. Participants may withdraw from the platform and discontinue data sharing at any time, retaining ongoing control over their contributed data. For any participants under the age of 16 years, a parent or guardian was required to register and complete the questionnaire on their behalf.

STW obtained ethics approval from the Helsinki/IRB Committee of Sheba Medical Center (Ramat Gan, Israel; protocol #7356-20-SMC) for the collection and analysis of de-identified data for medical research across its chronic disease communities. This study analyzed de-identified, routinely collected data from the STW digital health platform, presented to the authors in aggregate form. Given that each participant had previously consented at registration to the use of their de-identified data for research purposes, including by external researchers, and that secondary analysis of existing data posed minimal risk of re-identification—a key ethical consideration in digital health platform use [88]—the Sheba Medical Center Ethics Committee approved a waiver of requirement for additional consent for use of the medical questionnaire for research purposes. The present study falls within the scope of this approval.

Data collected by STW are stored in a de-identified manner, with each user assigned a non-indicative number. For the present study, data extraction and aggregation were performed within the STW platform by one author (ED) who was affiliated with STW at the time the data were accessed. The dataset provided to the remaining members of the research team contained only aggregated information summarizing symptoms, treatments, aggravating factors, and comorbidities at the population level. No individual-level data capable of identifying or re-identifying participants were shared with the authors. This study therefore represents secondary analysis of de-identified, aggregated user data. It was conducted in accordance with the principles of the Declaration of Helsinki and consistent with recognized ethical guidance for conducting health research with online communities [89].

## Results

### Demographics

Between July 2020 and December 2022, 1250 individuals enrolled on the STW platform as members of the arachnoiditis research community. Of these, 1105 (88.4%) affirmed that their diagnosis of arachnoiditis had been confirmed by a doctor, and only their data were included in the study cohort for the following analyses.

Study participants were international, with the majority (71.4%) from the United States, and three quarters (75.9%) were female. White or European ethnicities were predominant among survey participants, while black or African American, Asian, and Hispanic ethnicities were relatively underrepresented. Specific demographic characteristics of the study participants are summarized in Table 1.

**Table 1.** Demographic characteristics of 1105 study participants.

| Characteristic | n | % |
| --- | --- | --- |
| **Sex (at birth)** |  |  |
| Female | 839 | 75.9 |
| Male | 266 | 24.1 |
| **Country** |  |  |
| United States | 789 | 71.4 |
| United Kingdom | 77 | 7.0 |
| Canada | 50 | 4.5 |
| Australia | 31 | 2.8 |
| Germany | 12 | 1.1 |
| South Africa | 7 | 0.6 |
| Others | 139 | 12.6 |
| **Age at time of survey <sup>a</sup>** |  |  |
| 0-18 years | 3 | 0.3 |
| 19-30 years | 43 | 3.9 |
| 31-45 years | 252 | 22.8 |
| 46-65 years | 615 | 55.7 |
| 66+ years | 192 | 17.4 |
| **Age at time of symptom onset <sup>b</sup>** |  |  |
| 0-18 years | 26 | 2.5 |
| 19-30 years | 133 | 13.0 |
| 31-45 years | 389 | 38.1 |
| 46-65 years | 408 | 39.9 |
| 66+ years | 66 | 6.5 |
| **Ethnicity <sup>c</sup>** |  |  |
| White | 943 | 89.0 |
| European | 207 | 19.5 |
| American Indian or Alaska native | 45 | 4.2 |
| Hispanic or Latino | 45 | 4.2 |
| Asian | 20 | 1.9 |
| Black or African American | 16 | 1.5 |
| Arab or Middle East | 9 | 0.8 |
| **Body Build <sup>d</sup>** |  |  |
| Significantly underweight | 25 | 2.2 |
| Moderately underweight | 78 | 7.1 |
| Average weight | 441 | 39.9 |
| Moderately overweight | 452 | 40.9 |
| Significantly overweight | 109 | 9.9 |

*n*, number of participants in each category
<sup>a</sup> Percentages sum to 100.1% due to rounding
<sup>b</sup> Not all participants reported age at initial onset (*n* = 1022)
<sup>c</sup> Totals for ethnicity are greater than 1105, and percentages exceed 100%, as participants were able to select multiple responses for ethnicity, as applicable
<sup>d</sup> Body build was self-reported by participants

Participant age at the time of taking the survey and at the appearance of initial symptoms are presented in Fig. 1. The majority of respondents at the time of the survey (73.1%) were ≥ 46 years old. At the time of initial symptom onset there is a proportional shift to the younger ages, with more than half (53.6%) ≤ 45 years.

**Fig. 1.**
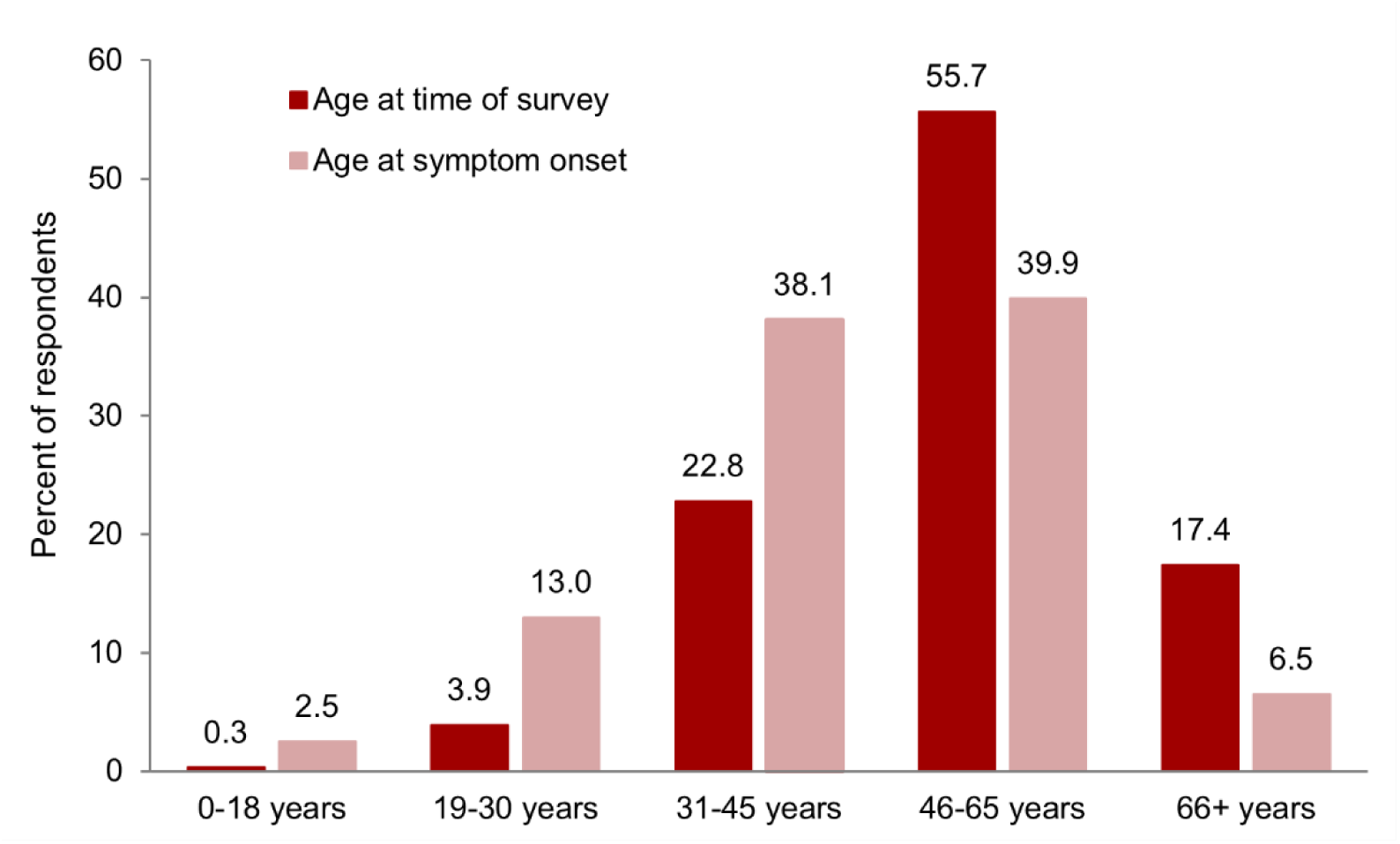
Comparison of age distribution at time of survey and at onset of symptoms among 1105 participants. Not all participants reported age at symptom onset (n = 1022).

### Assessment of symptoms

Participants were asked to list their symptoms in order of severity. Almost all participants (99.6%, n = 1101) responded to this question, including 74 respondents who listed no specific symptoms. A total of 712 specific symptoms were described. Fig. 2 displays the 20 most frequently reported symptoms and their percentages, with color coding to denote symptoms with STW cross-condition scores ≥ 6 (most indicative for arachnoiditis).

**Fig. 2.**
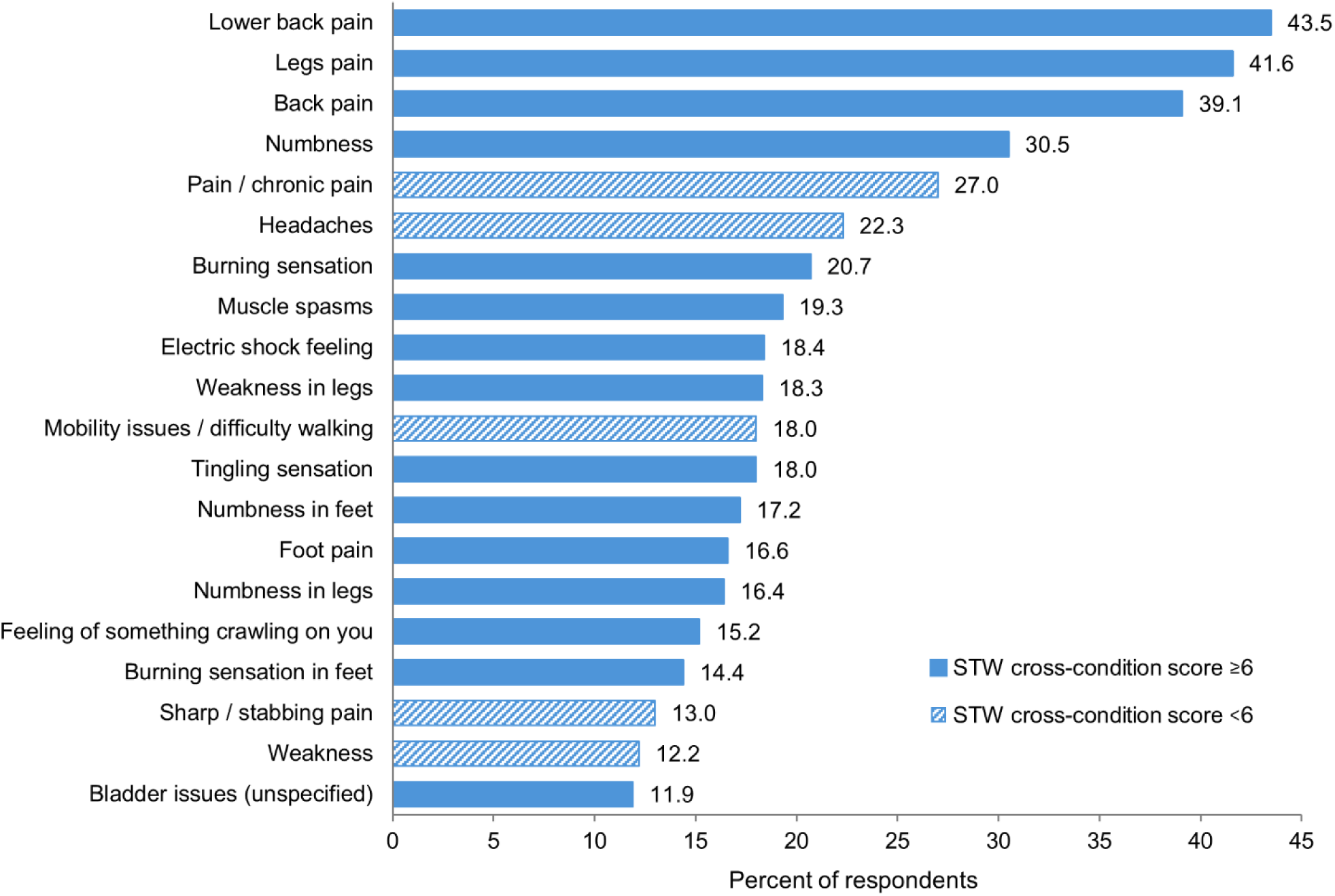
Most common symptoms among 1101 study participants reporting symptom data. STW (StuffThatWorks) cross-condition score (scale 1-10): A score ≥ 6 is highly indicative for arachnoiditis, a score between 3-5 has a medium degree of indication, and a score < 3 has a low indication (see Methods). Solid bars represent scores ≥ 6.

Given the large volume of symptoms described (n = 712), the 10% most frequently cited (n = 71) were categorized into groups of like symptoms using content analysis methodology. This produced 8 main symptom categories: pain; symptoms of motor nerve damage or irritation; altered sensation; symptoms experienced in the feet; bladder or bowel issues; head and neck symptoms; difficulty with daily activities; and “other” (autonomic, gastrointestinal, endocrine, autoimmune, heat or temperature sensation, and not otherwise categorized). Analysis of the additional 67 symptoms reported by ≥ 1% of the total study population (i.e. ≥ 11 people) did not yield any further distinct symptom categories beyond those previously identified.

Table 2 presents the symptoms in each category and corresponding percentages. STW cross-condition scores (also shown for each symptom), revealed a hierarchy of symptoms highly indicative of arachnoiditis (scores ≥ 6), with decreasing significance as follows: lower back pain, legs pain, formication (feeling of something crawling on you), electric shock feeling, burning sensation in feet, weakness in legs, numbness in feet, bladder issues, numbness in legs, burning sensation in legs, numbness, back pain, muscle spasms, foot pain, burning sensation, buttocks pain, shooting leg pain, and tingling sensation. Supplementary Material 2 provides a complete list of all symptoms reported by participants, ordered by frequency (n, %, STW cross-condition score).

**Table 2:** Main symptoms by category.

| Categories | Symptoms |  |  |  |  |  |
| --- | --- | --- | --- | --- | --- | --- |
|  | Percent of participants (n=1101); STW cross-condition score<sup>a</sup> |  |  |  |  |  |
|  |  | % | score |  | % | score |
| **Pain** |  |  |  |  |  |  |
| Spinal pain | Lower back pain | 43.5 | **7.21** | Back pain | 39.1 | **6.86** |
|  | Spine pain | 3.3 | 5.04 | Lumbar region pain | 3.1 | 5.65 |
|  | Pain across sacrum | 2.9 | 4.99 |  |  |  |
| Referred pain | Legs pain | 41.6 | **7.12** | Burning sensation in legs | 10.8 | **7.00** |
|  | Hip pain | 9.6 | 5.59 | Buttocks pain | 8.7 | **6.24** |
|  | Sciatica | 5.9 | 5.44 | Shooting leg pain / radiating pain down leg | 4.6 | **6.17** |
|  | Nerve pain in legs | 4.5 | 5.25 | Radiating pain | 4.2 | 5.00 |
|  | Right leg pain | 3.5 | 5.18 | Left leg pain | 2.9 | 5.24 |
|  | Groin pain | 2.9 | 3.80 |  |  |  |
| Pain quality | Burning sensation | 20.7 | **6.34** | Electric shock feeling | 18.4 | **7.08** |
|  | Sharp / stabbing pain | 13.0 | 5.93 | Nerve pain | 8.4 | 5.41 |
|  | Burning pain | 7.6 | 5.93 | Shooting pains | 6.7 | 5.61 |
|  | Stinging | 4.9 | 5.21 | Severe pain | 4.9 | 3.08 |
|  | Crippling / extreme pain | 3.7 | 5.11 |  |  |  |
| Pain other | Pain / chronic pain | 27.0 | 3.70 | Widespread pain / body aches | 3.5 | 2.73 |
| **Motor nerve damage or irritation** | Muscle spasms | 19.3 | **6.64** | Weakness in legs | 18.3 | **7.01** |
|  | Weakness | 12.2 | 5.01 | Foot drop | 5.7 | 5.54 |
|  | Neuropathy | 5.5 | 5.38 | Leg cramps | 5.1 | 5.23 |
|  | Twitching | 4.8 | 5.08 | Cramping | 4.6 | 3.14 |
|  | Spasms | 3.7 | 3.96 | Leg/s spasms | 3.4 | 4.88 |
| **Altered sensation** | Numbness | 30.5 | **6.96** | Tingling sensation | 18.0 | **6.05** |
|  | Numbness in legs | 16.4 | **7.00** | Formication (feeling of something crawling on you) | 15.2 | **7.09** |
|  | Tingling legs | 7.9 | 5.50 | Pins and needles | 5.4 | 4.79 |
|  | Lower leg numbness | 4.3 | 4.74 | Water dripping sensation | 3.8 | 5.63 |
| **Symptoms in feet** | Numbness in feet | 17.2 | **7.01** | Foot pain | 16.6 | **6.52** |
|  | Burning sensation in feet | 14.4 | **7.04** | Tingling feet | 6.0 | 5.06 |
| **Bladder or bowel symptoms** | Bladder issues (unspecified) | 11.9 | **7.01** | Incontinence | 10.3 | 5.69 |
|  | Bowel issues (unspecified) | 6.6 | 5.20 | Fecal incontinence | 4.2 | 5.14 |
| **Head and neck symptoms** | Headaches | 22.3 | 4.54 | Blurred vision | 8.0 | 5.21 |
|  | Balance issues / loss of balance | 7.5 | 4.83 | Neck pain | 6.6 | 4.37 |
|  | Migraines | 6.4 | 3.32 | Tinnitus | 6.2 | 4.96 |
|  | Dizziness / severe dizziness | 4.4 | 2.32 | Brain fog | 3.3 | 2.09 |
| **Difficulty with daily activities** | Mobility issues / difficulty walking | 18.0 | 5.48 | Difficulty standing | 9.4 | 5.82 |
|  | Difficulty sitting | 7.6 | 5.61 | Pain sitting | 3.8 | 5.44 |
|  | Unable to walk / stand | 3.0 | 3.72 |  |  |  |
| **Other** | Fatigue | 10.9 | 1.95 | Insomnia | 4.8 | 2.54 |
|  | Sweating / overheating | 4.5 | 3.55 | Depression / feeling depressed | 3.9 | 2.04 |
|  | Nausea | 3.1 | 1.72 |  |  |  |

*n*, number of participants who reported symptom data (1101); *STW*, StuffThatWorks
<sup>a</sup> STW cross-condition score (scale 1-10): A score ≥ 6 is highly indicative for arachnoiditis, a score between 3-5 has a medium
degree of indication, and a score < 3 has a low indication (see Methods). Scores ≥ 6 are shown in bold

### Earliest presenting symptoms

Participants were asked to recall whether, prior to diagnosis or early in the condition, there were any symptoms or warning signs that worried them or made them feel something was not right. The majority of participants (92.2%, n = 1019) endorsed early symptoms, reporting 418 symptoms, similar in character to the general arachnoiditis symptoms participants had listed. Pain and altered sensation were predominant among the most common reported early symptoms (14/20 symptoms). An absence of early symptoms was indicated by 8.8% of respondents. The frequency, percent and STW cross-condition score for the 20 most common early symptoms are presented in Table 3. Supplementary Material 3 provides a complete list of all early symptoms reported by participants, ordered by frequency (n, %, STW cross-condition score).

**Table 3.** Most common early symptoms among 1019 study participants reporting early symptoms.

| Early symptom | n | % | STW cross-condition score<sup>a</sup> |
| --- | --- | --- | --- |
| Pain / chronic pain | 172 | 16.8 | 5.40 |
| Back pain | 110 | 10.8 | **7.03** |
| None | 90 | 8.8 | - |
| Legs pain | 73 | 7.1 | **7.00** |
| Lower back pain | 51 | 5.0 | **7.00** |
| Mobility issues / difficulty walking | 48 | 4.7 | 5.17 |
| Numbness | 47 | 4.6 | **6.02** |
| Numbness in legs | 44 | 4.3 | **6.72** |
| Constant pain | 41 | 4.0 | 5.26 |
| Severe pain | 41 | 4.0 | 5.07 |
| Burning sensation | 39 | 3.8 | 5.79 |
| Weakness in legs / jelly legs | 36 | 3.5 | **6.07** |
| Numbness in feet | 34 | 3.3 | **6.16** |
| Deterioration of health / condition / worsening of symptoms | 31 | 3.0 | 2.82 |
| Electrical shock feeling | 30 | 2.9 | **6.10** |
| Burning sensation in legs | 28 | 2.7 | 5.92 |
| Surgery induced condition | 28 | 2.7 | 5.62 |
| Crippling / extreme pain | 27 | 2.6 | 5.58 |
| Headaches | 27 | 2.6 | 3.67 |
| Tingling sensation | 26 | 2.5 | 5.12 |

*n*, number of participants with each symptom; STW, StuffThatWorks.
<sup>a</sup> STW cross-condition score (scale 1-10): A score $\geq 6$ is highly indicative for arachnoiditis, a score between 3-5 has a medium degree of indication, and a score $< 3$ has a low indication (see Methods). Scores $\geq 6$ are shown in bold.

### Assessment of aggravating factors

Factors which exacerbated their condition were reported by 95.0% of respondents (n = 1050), comprising 276 different aggravating factors. The 20 most frequently reported aggravating factors are presented in Fig. 3. “Sitting or sitting too long” and “standing or standing too long” were the most common aggravating factors, reported by 62.5% and 58.3% of respondents, respectively, and carried the highest cross-condition scores for aggravating factors at 7.70 and 7.59, respectively.

**Fig. 3.**
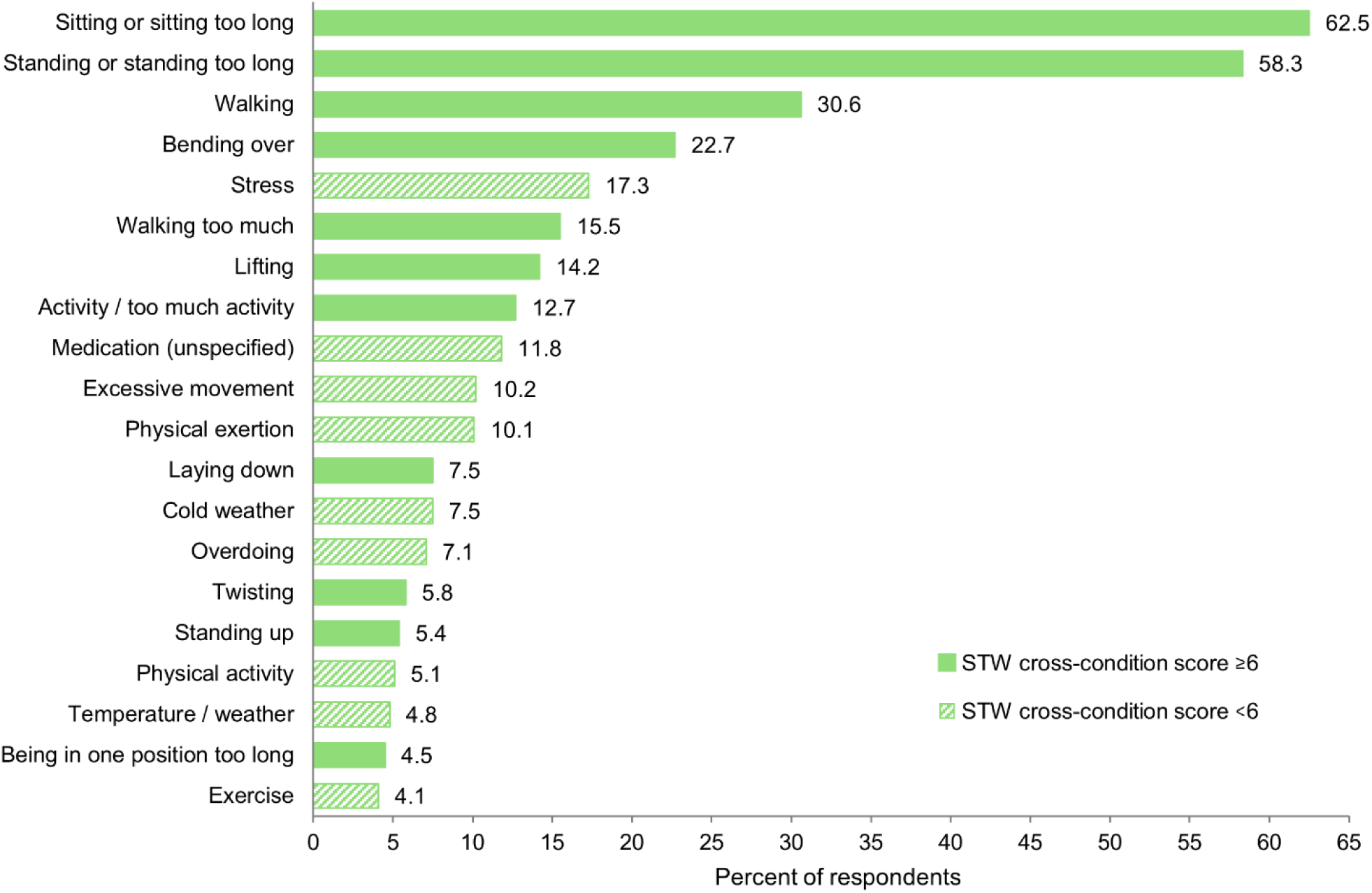
Most common aggravating factors among 1050 study participants reporting aggravating factors. STW (StuffThatWorks) cross-condition score (scale 1-10): A score ≥ 6 is highly indicative for arachnoiditis, a score between 3-5 has a medium degree of indication, and a score < 3 has a low indication (see Methods). Solid bars represent scores ≥ 6.

Analysis revealed 62 exacerbating factors reported by ≥ 1% of participants, stratified into eight discrete categories: positional factors (sitting, standing, or lying down); specific movements or activities; physical exertion intensity; weather or temperature variables; physical or mental health parameters; medical treatment or non-treatment; inactivity; and miscellaneous variables. Table 4 presents these factors by category, including their frequency and STW cross-condition scores. Supplementary Material 4 provides a complete list of all aggravating factors reported by participants, ordered by frequency (n, %, STW cross condition score).

**Table 4.**
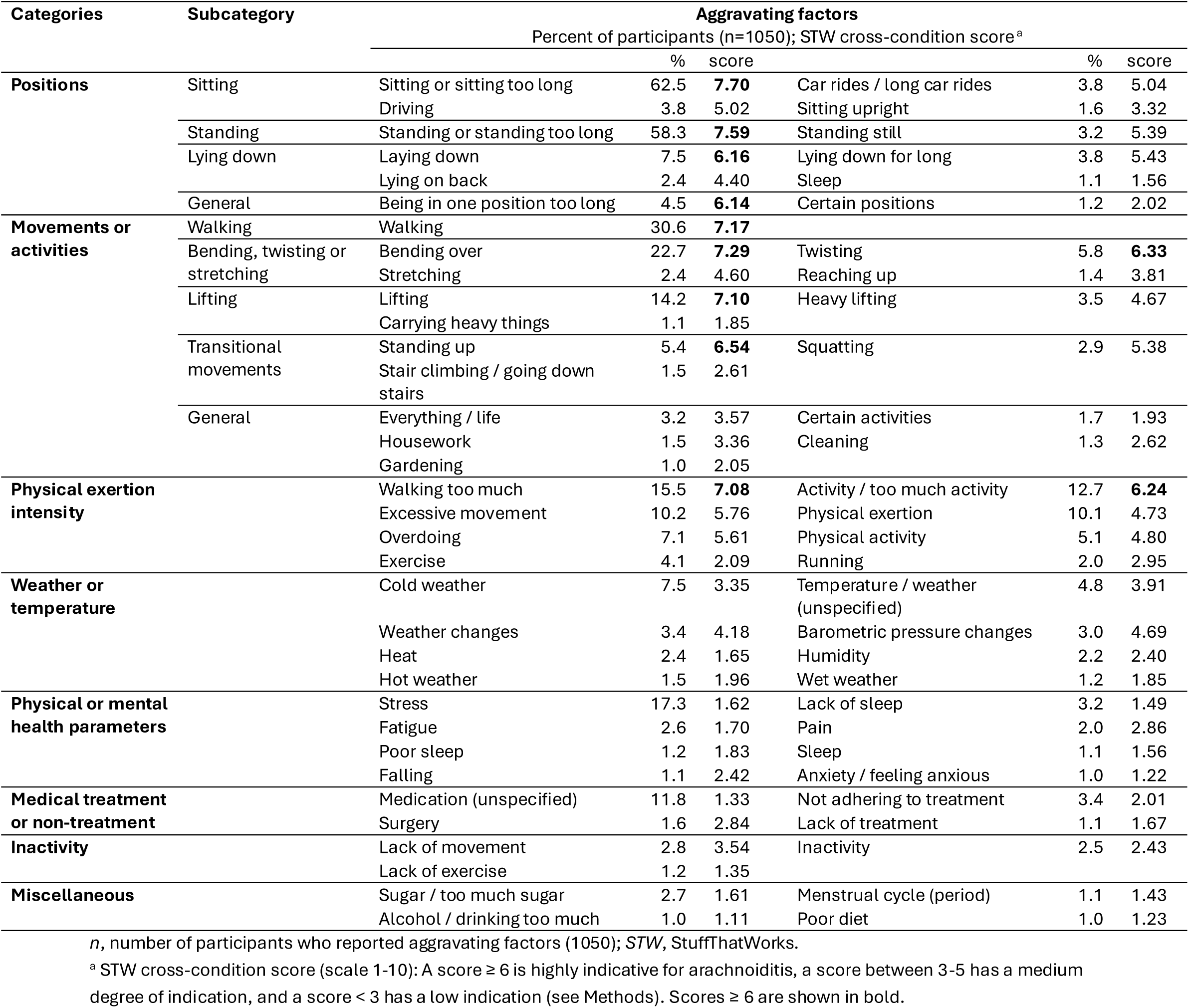
Aggravating factors by category.

### Assessment of comorbid conditions

Participants were asked to list all diagnosed comorbid medical conditions, with 830 (72.2%) respondents reporting at least one such condition. The 20 most common conditions are shown in Table 5 along with their frequency, % and STW cross-condition score. Eight comorbidities were spine-related in origin, seven of which yielded cross-condition scores highly indicative of arachnoiditis.

**Table 5.** Most common comorbidities among 830 study participants reporting comorbidities.

| Comorbidity | n | % | STW cross-condition score<sup>a</sup> |
| --- | --- | --- | --- |
| Degenerative Disc Disease (DDD) | 268 | 32.3 | **7.13** |
| Spinal stenosis | 210 | 25.3 | **7.33** |
| Fibromyalgia | 207 | 25.0 | 3.82 |
| Hypertension | 112 | 13.5 | 2.10 |
| Osteoarthritis | 103 | 12.4 | 4.97 |
| Arthritis (unspecified) | 99 | 11.9 | 5.04 |
| Cauda equina syndrome | 91 | 11.0 | **7.52** |
| Scoliosis | 85 | 10.3 | 5.86 |
| Clinical depression | 84 | 10.1 | 1.24 |
| Migraine | 83 | 10.0 | 2.03 |
| Failed back surgery syndrome (FBSS) | 82 | 9.9 | **7.43** |
| Tarlov cysts | 81 | 9.8 | **7.39** |
| Herniated disc | 70 | 8.4 | **7.02** |
| Hypothyroidism | 69 | 8.3 | 1.72 |
| Anxiety / generalized anxiety disorder (GAD) | 68 | 8.2 | 1.15 |
| Ehlers Danlos Syndrome (EDS) | 66 | 8.0 | 4.09 |
| Diabetes mellitus | 56 | 6.8 | 2.55 |
| Bulging discs | 53 | 6.4 | **7.00** |
| Complex regional pain syndrome | 50 | 6.0 | **6.43** |
| Rheumatoid arthritis | 47 | 5.7 | 2.93 |

*n*, number of participants with each comorbidity; STW, StuffThatWorks
<sup>a</sup> STW cross-condition score (scale 1-10): A score $\geq 6$ is highly indicative for arachnoiditis, a score between 3-5 has a medium degree of indication, and a score $< 3$ has a low indication (see Methods). Scores $\geq 6$ are shown in bold

### Survey of treatments

The 30 treatments most frequently tried by the 982 participants (88.9%) who responded to this question are shown in Table 6, ordered by self-rated effectiveness (STW estimated ratio). The 90% confidence intervals for rates of effectiveness are generally quite broad, reflecting the relatively small numbers of participants who rated individual treatments as effective.

**Table 6.** Most frequently tried treatments among 982 study participants reporting treatment data, ranked by self-rated effectiveness.

| Treatment tried | Total<sup>a</sup><br>n (%) | Self-rated as effective<sup>b</sup> |  |  |
| --- | --- | --- | --- | --- |
|  |  | n | STW estimated<br>ratio (%)<sup>c</sup> | 90% CI |
| Naltrexone hydrochloride (low dose naltrexone) | 67 (6.8) | 20 | 28.1 | 20.0-37.0 |
| Ketamine hydrochloride (ketamine infusions) | 65 (6.6) | 17 | 24.8 | 16.9-33.4 |
| Fentanyl (patch and other routes) | 92 (9.4) | 20 | 21.1 | 14.7-28.1 |
| Oxycodone hydrochloride | 231 (23.5) | 49 | 20.9 | 16.8-25.4 |
| Pain relief medication (unspecified) | 97 (9.9) | 20 | 20.1 | 14.0-26.8 |
| Morphine sulfate | 104 (10.6) | 21 | 19.7 | 13.9-26.2 |
| Spinal cord stimulator (SCS) | 119 (12.1) | 24 | 19.7 | 14.2-25.8 |
| Tizanidine hydrochloride | 77 (7.8) | 15 | 18.9 | 12.4-26.3 |
| Hydrocodone bitartrate | 72 (7.3) | 12 | 16.3 | 10.1-23.5 |
| Stretching | 72 (7.3) | 11 | 15.1 | 9.0-22.1 |
| Magnesium (glycinate and others) | 67 (6.8) | 10 | 14.7 | 8.6-21.9 |
| Paracetamol + oxycodone hydrochloride | 64 (6.5) | 9 | 14.0 | 7.9-21.2 |
| Pregabalin | 260 (26.5) | 36 | 13.8 | 10.5-17.4 |
| Tramadol hydrochloride | 109 (11.1) | 14 | 12.9 | 8.2-18.3 |
| Cannabidiol (CBD) | 79 (8.04) | 10 | 12.7 | 7.4-19.0 |
| Muscle relaxants | 65 (6.5) | 8 | 12.4 | 6.7-19.2 |
| Hydrotherapy | 76 (7.7) | 9 | 12.0 | 6.7-18.2 |
| Massage | 94 (9.6) | 11 | 11.8 | 7.0-17.4 |
| Chiropractic | 65 (6.6) | 7 | 11.0 | 5.7-17.6 |
| Paracetamol + hydrocodone bitartrate | 68 (6.9%) | 7 | 10.6 | 5.4-16.9 |
| Duloxetine hydrochloride | 135 (13.7) | 14 | 10.5 | 6.6-15.0 |
| Baclofen / baclofen intrathecal pump | 92 (9.4) | 9 | 10.0 | 5.6-15.4 |
| Gabapentin | 372 (37.9) | 37 | 10.0 | 7.6-12.7 |
| Turmeric capsules / pills | 74 (7.5) | 7 | 9.8 | 5.0-15.7 |
| Paracetamol (acetaminophen) | 76 (7.7) | 6 | 8.4 | 4.0-13.8 |
| Physiotherapy | 296 (30.1) | 21 | 7.2 | 5.0-9.8 |
| Amitriptyline hydrochloride | 81 (8.2) | 5 | 6.8 | 3.0-11.6 |
| Ibuprofen | 88 (9.0) | 4 | 5.2 | 2.1-9.4 |
| Transcutaneous electrical nerve stimulation | 85 (8.7) | 3 | 4.3 | 1.5-8.2 |
| Acupuncture | 90 (9.2) | 2 | 3.1 | 0.9-6.4 |

*n*, number of participants reporting each treatment; *STW*, StuffThatWorks; *CI*, confidence intervals
<sup>a</sup> Tried treatments were 90% normalized by the STW normalization tool (see Methods)
<sup>b</sup> Self-rated effectiveness = “significant improvement of condition”
<sup>c</sup> STW estimated ratio = binomial proportion with 90% confidence intervals (see Methods)

Among the treatments in Table 6, it can be seen that the most frequently tried treatments are not the same as those self-rated as most effective. The most frequently tried treatments (listed by ≥ 20% of respondents) include gabapentin (37.9%), physiotherapy (30.1%), pregabalin (26.5%), and oxycodone (23.5%). In contrast, the treatments with the highest rated effectiveness (≥ 20% estimated ratios) include low-dose naltrexone (28% effective, tried by only 6.8%), ketamine infusion (24.8%), fentanyl (21.1%), and oxycodone (20.9%).

The 30 treatments rated as most detrimental by participants are shown in Table 7. The most detrimental (≥ 20% estimated ratios) are: epidural steroid injections (38.5%), spinal injections (unspecified) (31.1%), myelogram (28.5%), epidural injections (unspecified) (28.2%), surgery (21.7%), and traction (20.4%).

**Table 7.**
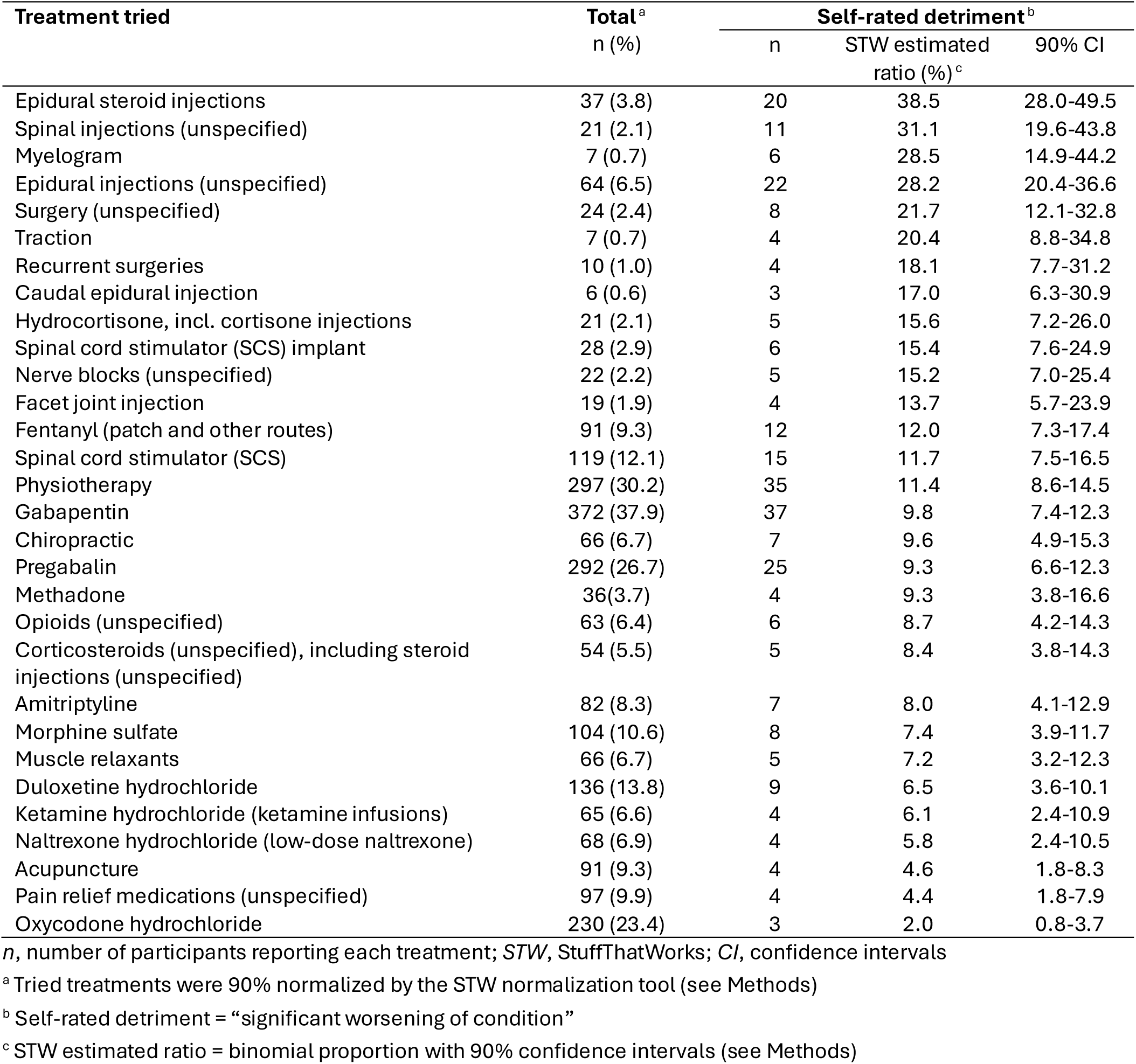
Most detrimental treatments among 982 study participants reporting treatment data, ranked by self-rated detriment.

| Treatment tried | Total<sup>a</sup><br>n (%) | Self-rated detriment<sup>b</sup> |  |  |
| --- | --- | --- | --- | --- |
|  |  | n | STW estimated<br>ratio (%)<sup>c</sup> | 90% CI |
| Epidural steroid injections | 37 (3.8) | 20 | 38.5 | 28.0-49.5 |
| Spinal injections (unspecified) | 21 (2.1) | 11 | 31.1 | 19.6-43.8 |
| Myelogram | 7 (0.7) | 6 | 28.5 | 14.9-44.2 |
| Epidural injections (unspecified) | 64 (6.5) | 22 | 28.2 | 20.4-36.6 |
| Surgery (unspecified) | 24 (2.4) | 8 | 21.7 | 12.1-32.8 |
| Traction | 7 (0.7) | 4 | 20.4 | 8.8-34.8 |
| Recurrent surgeries | 10 (1.0) | 4 | 18.1 | 7.7-31.2 |
| Caudal epidural injection | 6 (0.6) | 3 | 17.0 | 6.3-30.9 |
| Hydrocortisone, incl. cortisone injections | 21 (2.1) | 5 | 15.6 | 7.2-26.0 |
| Spinal cord stimulator (SCS) implant | 28 (2.9) | 6 | 15.4 | 7.6-24.9 |
| Nerve blocks (unspecified) | 22 (2.2) | 5 | 15.2 | 7.0-25.4 |
| Facet joint injection | 19 (1.9) | 4 | 13.7 | 5.7-23.9 |
| Fentanyl (patch and other routes) | 91 (9.3) | 12 | 12.0 | 7.3-17.4 |
| Spinal cord stimulator (SCS) | 119 (12.1) | 15 | 11.7 | 7.5-16.5 |
| Physiotherapy | 297 (30.2) | 35 | 11.4 | 8.6-14.5 |
| Gabapentin | 372 (37.9) | 37 | 9.8 | 7.4-12.3 |
| Chiropractic | 66 (6.7) | 7 | 9.6 | 4.9-15.3 |
| Pregabalin | 292 (26.7) | 25 | 9.3 | 6.6-12.3 |
| Methadone | 36(3.7) | 4 | 9.3 | 3.8-16.6 |
| Opioids (unspecified) | 63 (6.4) | 6 | 8.7 | 4.2-14.3 |
| Corticosteroids (unspecified), including steroid injections (unspecified) | 54 (5.5) | 5 | 8.4 | 3.8-14.3 |
| Amitriptyline | 82 (8.3) | 7 | 8.0 | 4.1-12.9 |
| Morphine sulfate | 104 (10.6) | 8 | 7.4 | 3.9-11.7 |
| Muscle relaxants | 66 (6.7) | 5 | 7.2 | 3.2-12.3 |
| Duloxetine hydrochloride | 136 (13.8) | 9 | 6.5 | 3.6-10.1 |
| Ketamine hydrochloride (ketamine infusions) | 65 (6.6) | 4 | 6.1 | 2.4-10.9 |
| Naltrexone hydrochloride (low-dose naltrexone) | 68 (6.9) | 4 | 5.8 | 2.4-10.5 |
| Acupuncture | 91 (9.3) | 4 | 4.6 | 1.8-8.3 |
| Pain relief medications (unspecified) | 97 (9.9) | 4 | 4.4 | 1.8-7.9 |
| Oxycodone hydrochloride | 230 (23.4) | 3 | 2.0 | 0.8-3.7 |

*n*, number of participants reporting each treatment; *STW*, StuffThatWorks; *CI*, confidence intervals
<sup>a</sup> Tried treatments were 90% normalized by the STW normalization tool (see Methods)
<sup>b</sup> Self-rated detriment = “significant worsening of condition”
<sup>c</sup> STW estimated ratio = binomial proportion with 90% confidence intervals (see Methods)

Comparing Tables 6 and 7, it is evident that for several of the interventions that appear on both lists, reports of an effective versus detrimental effect registered similar numbers. For example, 37 respondents reported that gabapentin was effective, while an equal number reported that it was detrimental.

## Discussion

This preliminary investigation represents the most comprehensive observational cross-sectional study of arachnoiditis documented to date and establishes the foundation for subsequent analytical frameworks. The patient-centered methodological approach enabled the assembly of a substantially larger cohort than conventional methods would allow. Our research not only consolidates what is already known about arachnoiditis but also presents several new findings.

### Prevalence according to demographics

Female participants comprised 75.9% of respondents in our study. This compares with 45.5% in a 2024 systematic review of 147 studies (510 cases) [7], suggesting a more balanced sex distribution in the broader literature. Other large (n ≥ 80) case series not covered in the systematic review have shown variable sex distributions, with female proportions ranging from 33% to 81% [4,12,28,34].

The true incidence of arachnoiditis and its sex ratio have not been well defined and may depend on causal factors, highlighting a need for further epidemiological studies. The marked female predominance seen in our study may reflect multiple factors. Autoimmunity may play a role in the pathophysiology of arachnoiditis, and the majority of autoimmune conditions are more common in females [90,91]. Another risk factor specific to females is obstetric epidural anesthesia. When carried out properly, epidural anesthesia is an extremely rare cause of arachnoiditis [30,92,93] however as up to two-thirds of females in developed countries undergo this procedure during childbirth [94], the sheer volume of cases augments the prevalence of this complication in proportion to the substantial number of procedures performed. It is also possible that one of the contributors to sex ratio imbalance in our study could be related to participation on the STW platform facilitated through social networks, in which females tend to be more engaged [95,96], although participants joined through multiple pathways.

In terms of age distribution, 73.1% of our respondents were ≥ 46 years old, which is older than the mean age of 44.2 years (standard deviation [SD] 17.53) reported in the 2024 systematic review [7]. This difference likely reflects our cross-sectional design, which captures individuals with established arachnoiditis rather than those at initial diagnosis. However, our participants’ ages fall within the SD of the review findings [7], and align with the aforementioned case series, which report median or mean ages of 48.4 to 61.3 years [4,12,28,34]. Importantly, our work adds insight as to the age of initial symptom onset, which is spread across earlier age groups, with 53.6% ≤ 45 years.

### Symptoms of arachnoiditis

The arachnoiditis symptoms we report are consistent with previous studies [1,4,10,12,34], however, our work further elaborates symptom descriptions, and also identifies which symptoms are most indicative of the condition. Pain and sensory disturbances were predominant among the most frequently reported symptoms (35/71 symptoms), and included burning sensations, electric shock sensations, shooting pain, numbness, tingling, and formication (sensations of insects crawling on the skin). All these symptoms had high STW cross-conditions scores, suggesting greater specificity for arachnoiditis. Formication has been described in previous reports [12,34] although it is not unique to arachnoiditis, and has also been reported with other neurological conditions [97]. Aldrete [12] has proposed neurophysiological explanations for many of the sensory symptoms in arachnoiditis, including formication, and also described pain distribution patterns that may not always follow nerve dermatomes but may be more regional or patchy in nature, reminiscent of findings in tethered cord syndrome [98,99].

Beyond sensory symptoms, signs of motor nerve damage or irritation are also frequently reported in our study, including weakness, foot drop, muscle spasms, cramps and twitching. Muscle related symptoms such as spasms, cramps and fasciculations have been previously reported [1,12] however they lack detailed characterization in the literature, warranting more comprehensive study to better understand their pathophysiology, temporal progression and their relationship to the location of spinal cord involvement. Motor symptoms suggest that electrodiagnostic tests may be useful, however, it has been noted that such tests do not produce a consistent pattern that can be identified as arachnoiditis for diagnostic purposes [3,4,12]. Yet, the potential utility of these tests has not been systematically investigated in individuals with arachnoiditis, particularly among those with a spectrum of motor nerve dysfunction, as has been studied in related conditions, such as adult tethered cord [100], syringomyelia [101], and Tarlov cysts [102,103]. Such research could prove beneficial for diagnosis, especially in suspected cases, as well as for monitoring disease progression.

An array of other physical signs and symptoms were described in people living with arachnoiditis in our cohort. Symptoms localized to the feet were common, with 17.2% reporting numbness, 16.6% pain, and 14.4% burning sensation, all of which were associated with high STW cross-condition scores ≥ 6. Exclusion of the more common causes of sensory dysesthesias in the feet seen in other conditions, along with the presence of other characteristic findings and risk factors consistent with arachnoiditis, may lead to more accurate diagnosis and shorten time to diagnosis.

Eight diverse types of head and neck symptoms were reported by study participants, led by headache (22.3%), however none produced a STW cross-condition score indicating a high specificity for arachnoiditis. Bladder and bowel symptoms, led by “unspecified bladder issues” (11.9%) and incontinence (10.3%), have also been reported in many previous studies [1,12,34,47] with only “unspecified bladder symptoms” producing a cross-condition score ≥ 6.

### Earliest presenting symptoms

We sought to characterize the earliest presenting symptoms of arachnoiditis given there is some evidence that disease progression might be arrested or reversed in some individuals with early intervention. Early recognition may also provide patients with timely explanations for their symptoms and help to reduce diagnostic uncertainty. Our preliminary results suggest that the character and pattern of earliest symptoms do not differ significantly from general reported symptoms. Pain and altered sensation predominated among the top 20 earliest symptoms. Future studies will benefit from gathering information about initial symptoms as soon as possible after diagnosis to gain a more accurate picture of symptom development.

### Nature of aggravating factors

The most frequently reported aggravating factors, “sitting or sitting too long” and “standing or standing too long”, were reported by 62.5% and 58.3% of the study population, respectively. The positional nature of these and other commonly reported aggravating factors is noteworthy, both in our study and in previous reports. Porcelli and Tennant [34] reported that 87.5% of their 80 subjects with lumbosacral arachnoiditis experienced relief of symptoms upon standing up, 86.3% reported that standing up for too long caused the need to lie down, and 83.8% of their subjects found that it hurt to lie flat on their back. Similar positional symptoms have been documented in case reports of arachnoiditis [18,104,105]. The specific positions that aggravate symptoms may be correlated with the spinal levels that are affected by arachnoiditis (not assessed in this study), such that the inflamed or adhered nerves are compressed or stretched in certain positions. In tethered cord syndrome, positional symptoms caused by elongation of the lumbar spine have been reported [98], and a potentially analogous mechanism may be relevant in some cases of arachnoiditis. Positional aggravating factors may also be reflective of their effect in increasing CSF pressure in various parts of the spine against an arachnoiditis-induced CSF flow disturbance, or they may be suggestive of a particular complication. In future studies, we intend to correlate the level of spinal involvement with specific aggravating factors.

Movements that provoked symptoms were categorized as walking, lifting, transitional movements, and activities that bend, twist or stretch. Symptom exacerbation likely involves multifactorial mechanisms. For example, when a nerve is inflamed or irritated, it becomes more sensitive to mechanical forces like stretching or compression [106]. Additionally, in disease conditions causing mechanical nerve compromise such as tethered cord syndrome or radiculopathy, stretching of nerve roots contributes to pathophysiology due to limitation in nerve root movement or temporary ischemic changes [50,107,108], a finding which has been extended to arachnoiditis [50].

Activities related to lifting may involve additional mechanisms. During lifting the core abdominal muscles are activated, and in heavy lifting a Valsalva maneuver may also occur. These muscle activations increase intra-abdominal and intra-thoracic pressure transiently [109], both of which increase CSF pressure or volume [110–113]. Such changes can alter CSF flow in individuals with arachnoiditis or increase pressure against the nerve roots, potentially exacerbating neurologic symptoms. Increases in CSF pressure have been associated with symptom exacerbation in individuals with related spinal conditions including syringomyelia [114] and Tarlov cysts [103]. Patients with syringomyelia are generally advised to avoid deadlifts, as the Valsalva maneuver can result in deleterious syrinx expansion [114].

Spinal comorbidities, which were common among study participants, present additional complexities when interpreting aggravating factors, as they may share some positional and movement-related triggers.

Intense physical exertion commonly caused exacerbation of symptoms. While research shows benefits of physical activity for chronic or neuropathic pain [115–118], low or low-moderate intensity exercise is typically recommended—aligning with our findings. Individually tailored recommendations for physical activity and activity modification in individuals with arachnoiditis are generally advised.

Some aggravating factors described in our study, while commonly reported, showed low STW cross-condition scores and can be seen across multiple conditions besides arachnoiditis. These factors include variations in the weather or temperature, poor sleep, stress, anxiety, and lack of movement.

Our analysis has the potential to facilitate the development of broader and more precise clinical diagnostic criteria for arachnoiditis based on symptoms and aggravating factors, improving diagnostic accuracy and differential diagnosis. This would be especially important for those in whom arachnoiditis is suspected, but there are no correlative signs evident in radiological studies. Furthermore, reducing or avoiding exposure to an individual’s known aggravating factors has practical relevance for symptom management. Since neuropathic pain in individuals with arachnoiditis is often delayed in onset and protracted in duration relative to the inciting activity [1] the empiric pacing of known aggravating activities requires anticipation to avoid triggering symptom flares.

### Comorbid conditions

Coexisting degenerative spine conditions or anatomic abnormalities are frequently listed as comorbidities in our study, including spinal stenosis, cauda equina syndrome, scoliosis, and disc-related pathologies such as degenerative disc disease, or herniated or bulging discs. All of these conditions, except scoliosis, had high STW cross-condition scores, suggesting they correlate strongly with arachnoiditis. While these conditions have been associated with lumbar arachnoiditis in many earlier reports [4,7,12,14,28], they also lead to invasive spinal interventions—a well described risk factor that is often concomitantly reported in arachnoiditis studies (not ascertained in this study). However, both disc pathology [29,104] and degenerative spinal conditions [28,39,49,119,120] have also been identified as the sole apparent risk factor in cases of arachnoiditis with no history of spinal interventions.

In a related finding, Parenti et al. [43] demonstrated that spinal comorbidities in lumbar arachnoiditis have a significant relationship to arachnoiditis symptom dynamics, suggesting that integrated treatment of these comorbidities may help mitigate symptom progression. However, treatment decisions must carefully consider the potential for invasive interventions to worsen arachnoiditis. Although Palackdharry et al. [1] reported finding no evidence that surgical procedures (or lumbar puncture) in unaffected adjacent spinal levels worsen existing arachnoiditis, this was reported without detail. Further studies are required.

Failed back surgery syndrome was listed by 10% of our study participants as a comorbidity, consistent with previous studies [1,12,14,121]. Failed back surgery syndrome is an umbrella term for a variety of pathological conditions, including arachnoiditis, that result in persistent symptoms after spine surgery. The extent to which symptomatic lumbosacral arachnoiditis may be present prior to spinal surgery is unknown. This may be difficult to ascertain due to the overlap of symptoms between arachnoiditis and the underlying pathology necessitating surgery. In their study of patients undergoing surgery for spinal stenosis—a condition that is both a known cause of arachnoiditis due to mechanical irritation and creates confounding radiological findings due to the narrowed spinal canal [119,120,122]—Kawauchi et al. performed pre-operative myeloscopic assessment of the subarachnoid space [71]. They found that all patients demonstrated some degree of pre-operative arachnoid adhesions, however those with marked pre-operative adhesions experienced worse outcomes after the spinal stenosis surgery. Where surgeons are interested in determining if arachnoiditis is present pre-operatively and radiological findings are ambiguous, the expanded symptoms list may assist, particularly when patients report additional symptoms more suggestive of arachnoiditis (for example, burning sensations in the feet, or formication) than those typically associated with their degenerative condition.

Our study revealed a 9.8% prevalence of Tarlov cysts in our sample, approximately twice the estimated prevalence in the general population [123], supporting the association of Tarlov cysts with arachnoiditis proposed in previous reports [34,37,38]. A further 8.0% of respondents reported a diagnosis of EDS, which is higher than the estimated prevalence of joint hypermobility spectrum disorders in the general population (3-4%), and considerably higher than typical rates of EDS [124], consistent with this association in earlier reports [34,35]. This association, however, was not explicitly noted in a 2017 comprehensive review of the neurological and spinal manifestations of EDS [125], suggesting it may be underrecognized. Fibromyalgia was reported as a comorbidity in 25.0% of study participants, but produced a low STW cross-condition score. Given that fibromyalgia is a diagnosis of exclusion, with some symptoms overlapping those attributed to arachnoiditis, fibromyalgia could be misdiagnosed in patients with arachnoiditis. However, fibromyalgia demonstrates similarly high comorbid rates across various chronic conditions, including inflammatory arthritis, autoimmune disorders, centralized pain syndromes and neurological diseases, where it typically adversely affects quality of life and health status in these individuals [126]. The substantial prevalence of fibromyalgia in our cohort suggests it may represent a genuine and clinically important comorbidity in arachnoiditis patients.

Depression and anxiety, commonly associated with chronic pain conditions, were also frequently reported as comorbidities in our study. Even though osteoarthritis, reported by 12.4% of respondents, is highly prevalent in the age-related general population and less indicative of arachnoiditis according to a low cross-condition score, it should be noted that people with arachnoiditis and syrinx may experience Charcot-type neuropathic arthropathy in a variety of joints [127]. Hypertension, another comorbidity with a low cross-condition score, is also prevalent in the general population. However, episodic or paroxysmal surges of severe hypertension may be a sign of autonomic dysreflexia in individuals with arachnoiditis or syrinx [65,128,129]. In future studies, gathering more detailed information regarding comorbidities will elucidate etiological factors and predisposing conditions.

### Tried treatments

Participants in our study attempted a wide range of treatments, including pharmaceutical analgesics and neuropathic pain-directed medications, muscle relaxants, interventional pain procedures, over-the-counter pain relievers, cannabinoids, natural supplements, bio-electrical stimulation treatments, off-label treatments, and physical therapies. This range highlights that current approaches are predominantly focused on symptom management rather than addressing underlying inflammatory or disease-modifying mechanisms.

Confirming the consensus from earlier literature reports [1,3,11,12], our study demonstrates that arachnoiditis remains challenging to treat. Participant-reported treatment effectiveness rates ranged from 3.1% to 28.1%, with most treatments showing less than 15% effectiveness, demonstrating the critical need for developing more effective treatment strategies for this condition. Importantly, the wide confidence intervals for effectiveness indicate considerable uncertainty in these estimates, meaning the true rates could be much lower or higher.

Among the treatments, it is notable that the most frequently tried treatments are not the same as those rated as most effective. The most commonly tried treatments—gabapentin (37.9% of participants), physiotherapy (30.1%), and pregabalin (26.5%)—showed relatively modest effectiveness rates of 10.0%, 7.2%, and 13.8%, respectively. This pattern likely reflects standard clinical practice, where these treatments are commonly prescribed as first-line therapies for neuropathic pain due to their established use and familiarity among clinicians. In contrast, several less frequently used treatments demonstrated higher effectiveness rates.

Low dose naltrexone emerged as the most effective participant-reported treatment (28.1% effectiveness) while being tried by only 6.8% of participants. Several recent reports propose naltrexone as a treatment option for arachnoiditis [1,11,34], however there has been an absence of studies evaluating it in arachnoiditis patients. The preliminary data from our study suggests that naltrexone offers beneficial effects, with few detrimental reports, making it an attractive candidate for further study.

Ketamine also showed high participant-reported effectiveness rates (24.8%) while being tried by only 6.6% of participants. Its limited use likely reflects significant practical constraints, including controlled substance regulations, requirements for specialized infusion facilities, and notable side effects. Many of the highest-rated treatments face similar implementation challenges. These constraints highlight the broader challenge in chronic pain management of balancing therapeutic efficacy with safety and regulatory concerns.

Spinal cord stimulators (SCS) showed a moderate participant-reported effectiveness rate of 19.7%, placing them among the more effective treatments within our study. However, a considerable proportion (11.7%) of individuals who reported using this modality also rated SCS as detrimental. SCS have been recommended as a treatment modality option for arachnoiditis [1], although it has been noted that higher effectiveness rates are typically observed among specially selected patient subgroups. Further investigation into the specific factors contributing to treatment detriment in individuals with arachnoiditis, including patient selection criteria and optimal timing of intervention, will be important for future research.

While non-pharmacologic therapies such as stretching, hydrotherapy, and massage showed only modest effectiveness rates in our study (15.1%, 12.0%, and 11.8%, respectively), their favorable safety profiles make them important considerations for patient care, particularly as components of multimodal treatment approaches. Importantly, our findings from the assessment of aggravating factors indicate that certain physical activities, including stretching and overexertion, can worsen symptoms in some patients. Therefore, careful monitoring and gradual, individualized implementation of these therapies is essential.

Beyond the commonly reported treatments in our results, many other medications and interventions that have been identified as potentially beneficial in recent arachnoiditis papers were not represented in sufficient numbers in our study population to appear in our results.

These include, but are not limited to, ketorolac [34], diclofenac and other non-steroidal anti-inflammatories [3,65], corticosteroids [11,34,104,130], electrotherapeutic agents [65], metformin [1], cerebrolysin [1], eicosapentaenoic acid (fish oil) [104], newer formulations of curcumins [1,65], cognitive behavioral therapy to help modulate pain perception [3], and neural mobilization [1,105]. Similarly, many authors recommend multimodal and interprofessional pain management strategies, however there were not any multi-drug regimens or combination therapies reported with sufficient frequency to appear in our results.

Notably, the prevalence of corticosteroid use may be underestimated in our sample as a consequence of steroid medications being listed individually (e.g., dexamethasone, methylprednisolone) and therefore counted individually rather than as a collective category. Further, the open-text survey questions did not differentiate between treatments for routine daily symptom management versus periodic symptom flares. Since corticosteroids are frequently used to treat symptom flares and reduce inflammation [11,34,65,130], participants may have considered only their routine treatments when responding.

Analysis of data comparing treatments rated as effective versus detrimental revealed a paradoxical pattern wherein certain interventions received contradictory evaluations from different respondents. This may reflect inter-individual physiological differences, variations in disease severity or stage at the time of treatment, therapeutic efficacy that diminishes over time, or differences in underlying etiology. Different causes may elicit distinct inflammatory patterns or therapeutic windows. In the current study, however, we were unable to compare the efficacy of treatments over time or across different etiologies.

The most detrimental treatments in our study primarily consisted of invasive spinal interventions, reflecting their shared capacity to induce spinal cord irritation and inflammation directly though physical or mechanical trauma to already compromised neural tissues. This finding aligns with existing literature [1] that identifies invasive spinal procedures as risk factors for arachnoiditis development.

### Contribution of artificial intelligence (AI)/machine learning

Natural language processing techniques, coupled with artificial intelligence and machine learning algorithms, facilitate the analysis of extensive datasets, and have the potential to generate valuable and unique insights. The findings from our study offer considerable new details about arachnoiditis and would not have been feasible without an international online platform and the novel AI tools employed. The STW cross-condition score is particularly unique in its ability to compare individual symptoms, comorbidities and aggravating factors across very large numbers of people with and without arachnoiditis on the online platform, with each receiving a score indicating its specificity for the condition.

AI-generated data output, however, requires careful review and interpretation. For example, in the present study, some similar or identical symptoms were tabulated by AI machine learning as separate entities, such as “tingling” and “pins and needles”. This may have resulted from an incomplete ability of natural language processing to detect various open-label text terms as being synonymous. Further training of AI may be required to better recognize and properly categorize and group terms. Use of AI and machine learning to process and categorize large datasets and images in future studies may help to further establish accurate diagnostic criteria for arachnoiditis, including improved MRI interpretation [131,132].

### Study limitations

The most notable study limitations include reliance on self-reported data with a potential for recall bias. Furthermore, while the use of open-text survey questions enables a wide range of responses to facilitate learning as much as possible about the condition in an unbiased manner, this approach may be associated with more unintentional omissions compared to selecting options from a checklist.

This study did not address the duration, frequency and severity of symptoms, including symptom flares, and therefore cannot provide an overview of how pain and other symptom patterns differ between participants. In addition, the aggregate nature of the data provided, combined with the absence of data on disease location, extent, or suspected causes, prevented analysis by these variables, which may have yielded valuable insights.

As a cross-sectional study, our data on treatment effectiveness do not offer the same level of evidence as controlled clinical trials, and the binary effectiveness measure (self- reported significant improvement of condition) may underestimate treatments that provide modest but clinically relevant symptom relief. However, our aggregated findings may reveal meaningful trends and identify treatments suitable for future studies or individual trials. Assessment of treatment effectiveness is also complicated by the poorly defined natural history of arachnoiditis, often punctuated by symptom flares, and in which some individuals experience disease progression, whereas others remain static [2,4]. Treatments that successfully prevent deterioration rather than provide improvement may represent meaningful clinical success in some patients but may not be assessed as such. In addition, given that our results revealed a wide variety of treatments being used, a further limitation is that our data for effectiveness is limited to the 30 most frequently reported treatments. As a consequence, less frequently used but potentially more effective treatments may not have been captured. Specific adverse events or detrimental responses that might have led to discontinuation of a given treatment were beyond the scope of this present study, as were questions that could link specific treatments or sequencing of treatments to the early or advanced stage of disease for an individual.

Generalizability of our findings may be limited given that participation on the STW platform was driven in part by social media engagement, which tends to favor females and those with higher socioeconomic backgrounds [133]. In a similar vein, the majority of our respondents were of white/European ethnicity, which may be related to the survey being available only in the English language, or potentially also to the lower rates of social media involvement among other ethnic groups [134]. Similarly, completion of online questionnaires requires digital literacy and stamina, which could prevent those with significant disability levels from responding. Despite the potential for selection bias and lack of full representation of the entire population of those living with arachnoiditis, our findings remain consistent with earlier reports.

### Future directions

Significant areas of unmet need include consensus development of diagnostic criteria to reduce delays in diagnosis, establishment of natural history studies to better delineate differences between individuals in initiation and progression of disease, and discovery and development of more effective treatments.

Greater awareness of arachnoiditis and its causes, along with the development of more explicit diagnostic criteria, will aid in accelerating diagnosis and reducing the diagnostic “odyssey” so common in rare diseases [135,136]. Such enhanced recognition by the broader medical community, and improved understanding of the complications, can bring about a three-fold benefit. First, to highlight the essential need for multi-disciplinary care; second, to stimulate pharmaceutical industry and stakeholder interest in initiating and funding therapeutic research; and third, importantly, to benefit those individuals living with arachnoiditis who are actively seeking care, to be heard and understood by health care providers. With more information and training, providers may be less likely to be dismissive of persons affected by the condition, whose disabling symptoms frequently remain externally invisible to others, or to misattribute these symptoms to mental health issues or drug-seeking behavior [1,12,137–139].

The development of novel, more effective treatments for arachnoiditis must be a high priority as the science and understanding of modulating neuropathic and centrally mediated pain advance [73]. Given the inflammatory origin of arachnoiditis, future studies might explore whether medications that target inflammation and effectively cross the blood-brain barrier might both relieve symptoms and modify disease progression [1,16,73,140]. Treatments that prevent, stop or reverse fibrotic transformation and its consequences should be a high priority [1]. Advancements in minimally invasive spinal surgery are promising and may offer new treatment avenues for arachnoiditis patients in the future, with reduced morbidity [1]. The development of validated quality-of-life measures specifically designed for assessing interventions, like those recently developed for Tarlov cyst surgery [141], would significantly enhance our ability to measure treatment outcomes.

Research should focus not only on developing more effective therapies, but also on identifying biomarkers or clinical predictors that can guide personalized treatment selection, particularly given the potential timing-dependent efficacy of some treatments, and the substantial inter-individual variation in treatment responses observed in our study. In one example of such translational research, Zhang et al. [16] recently delineated differential gene expression from multiple cell types in human arachnoid tissues obtained from patients with and without arachnoiditis undergoing neurosurgery for Chiari malformation or syringomyelia. They employed transcriptome profiling using sn-RNA seq analysis, providing an avenue of discovery for new insights into the pathophysiology and signaling pathways characterizing adhesive arachnoiditis, with the possibility of biomarker discovery.

To support such research endeavors, an arachnoiditis international patient registry would enable comprehensive data collection across diverse populations over time and facilitate cross-disciplinary research collaboration. Such a registry could inform future prospective study design by employing a relational database in which sequential treatments are linked with clinical disease stage and treatment outcomes. Quantification of treatment effects can be accomplished through incorporation of validated patient reported outcome instruments evaluating quality of life, pain, disability, and health-related economic costs. It would be advantageous to store select anonymized patient health data such as MRI, EMG and other reports to confirm diagnosis, clarify clinical findings and elaborate upon patient generated data. Serial collection of longitudinal data on symptoms and level of disability would help to better define the natural history of progression in our sample. This would also permit interrogation of the larger dataset to explore any correlations that may exist between the presumptive inciting causes of an individual’s arachnoiditis, the disease distribution within the spinal cord, and the stage and severity of the condition. Such an approach might also enable exploration of whether certain subsets of patients respond differentially to various treatments or experience different rates of disease progression.

## Conclusion

This report describes the largest reported observational study on arachnoiditis to date. Our findings consolidate existing knowledge about the condition while advancing new and important observations regarding age of first symptom onset, range of symptoms, aggravating factors, comorbid conditions and self-assessment of tried treatments. Through the application of novel AI/machine learning algorithms we generated unique insights into symptom profiles, treatment patterns and diagnostic indicators for arachnoiditis, offering clinicians practical tools to improve diagnosis and management.

This work establishes a novel methodological approach to assembling a large patient cohort for rare disease research—a substantially larger cohort than would be possible through conventional methods. This advocacy-enabled, patient-centered model illustrates how rare disease advocacy networks can enable the large-scale data collection necessary for meaningful discovery in a condition that remains understudied and underdiagnosed.

## Data Availability

All data produced in the present study are contained in the manuscript and linked supplemental files.

## Supplementary Information

**Supplementary Material 1.** The StuffThatWorks (STW) survey questions used for the data collection for this analysis (PDF file).

**Supplementary Material 2.** A complete list of all symptoms reported by study participants, ordered by frequency (XLXS file). Includes frequency (n), percent, and STW cross-condition scores.

**Supplementary Material 3.** A complete list of all early symptoms reported by study participants, ordered by frequency (XLXS file). It includes frequency (n), percent, and STW cross-condition scores.

**Supplementary Material 4.** A complete list of all aggravating factors reported by study participants, ordered by frequency (XLXS file). It includes frequency (n), percent, and STW cross-condition scores.

## Abbreviations

ACMCRN: Arachnoiditis and Chronic Meningitis Collaborative Research Network
AI: Artificial intelligence
CSF: Cerebrospinal fluid
EDS: Ehlers Danlos Syndrome
MRI: Magnetic resonance imaging
SCS: Spinal cord stimulator
STW: StuffThatWorks

## Declarations

### Ethics approval and consent to participate

This study involved secondary analysis of de-identified, aggregated data from the StuffThatWorks digital health platform. The use of these data for research was approved by the Helsinki/ Ethics Committee at Sheba Medical Center, Israel (protocol #7356-20-SMC). Before sharing any data, all participants provided digital consent upon platform registration by agreeing to the platform’s terms of use and privacy policy, which informed them that their de-identified data may be used for research purposes, including by external researchers. The approving ethics committee deemed this prospective consent adequate and approved a waiver of requirement for additional consent for use of data from its medical questionnaire for research purposes, further stipulating that any data shared with external researchers must be in aggregate form only. Further details are provided in the Methods section.

### Consent for publication

Not applicable. The manuscript does not contain any identifiable individual data.

### Availability of data and materials

All aggregated data generated or analyzed during this study are included in this published article and its supplementary information files.

### Author contributions

Conceptualization: LV, NM, MB, PG, LB. Data curation: ED, MB, SS, LB. Formal analysis: ED, MB, IS, LB. Investigation: LV, ED, PG. Methodology: LV, NM, ED, MB, LB. Resources: LV, DRB, SS. Supervision: LV, LB. Visualization: NM, MB, DRB, SS, LB. Writing – original draft: MB, LB, NM, RS. Writing – review & editing: LV, NM, ED, RS, MB, PG, DRB, SS, LB. All authors have read and approved the final manuscript.

## Acknowledgements

The authors wish to thank all the individuals living with arachnoiditis who participated on the STW platform. We acknowledge StuffThatWorks Ltd for use of their digital platform, data aggregation and insights generation. We are grateful to Eve Blackburn for her efforts in raising awareness of the STW platform among individuals with arachnoiditis. We thank Michele Hansen PhD MPH and Richard Dobson MD for reviewing the manuscript, and Rob Lloyd PhD for his review of the CSF dynamics content. We are grateful to Enoch Carter for assistance with data preparation. We appreciate the continuing graphic design support of Matthew Churchward and Eve Blackburn. We dedicate this paper to the memory of ACMCRN co-founder and former VP of Research, Terri Lewis, PhD.

