## Supplemental Material 1 for "Arachnoiditis: leveraging crowdsourcing and AI in a cross-sectional study of 1,105 cases to improve identification, understanding, and treatment"

### StuffThatWorks (STW) survey questions used in data collection

- How old are you?

- Your sex at birth:
  - Female
  - Male
  - Intersex

- Where do you live?
  City, Country OR State, Country

- Was your arachnoiditis diagnosed by a doctor?
  - Yes
  - No

- How much time has passed since your arachnoiditis symptoms started?
  Ex: 15 years / 7 months etc.

- List your symptoms before starting treatment, in order of severity:
  Separate symptoms with a comma.

- Were there any early symptoms / warning signs? If yes, please list them.
  Tell us what got you worried or made you feel something wasn't right.

- Have you been diagnosed with any other conditions?
  - Yes
  - No

- [If yes]: Please list these conditions below:

- List ALL the treatments you've tried to date for arachnoiditis:
  *** HIGHLY IMPORTANT QUESTION ***
  INCLUDE ANYTHING YOU'VE TRIED: medicines, supplements, procedures, protocols, dietary changes, therapies, classes... PLEASE BE SPECIFIC: For drugs & supplements - the name of the drug as written on the package. Dietary changes - write which changes. Therapy - what kind? Separate items with a comma.
  If you haven't started treatment yet, or are not taking any measures to treat your arachnoiditis, please type "none."

- Did one (or a combination of) treatments SIGNIFICANTLY IMPROVE your condition?
    - Yes
    - No
    - Not sure
    - I haven't started treatment yet
    - I'm not taking any measures to treat this condition

- [If yes]: Write the name of the treatment (or combination) that SIGNIFICANTLY IMPROVED your condition:
  *** HIGHLY IMPORTANT QUESTION ***
  As above, include everything in you think was responsible for the improvement and be specific.

- Did you experience a treatment (or combination) that had a detrimental impact on your condition?
    - Yes
    - No
    - Not sure

- [If yes]: What was this detrimental treatment (or combination)?
  *** HIGHLY IMPORTANT QUESTION ***
  As above, include everything you think was responsible for the detrimental impact and be specific.

- To your knowledge, is there anything that makes your condition worse?
  Separate items with a comma, or simply write "No."

- How would you describe your weight?
    - Significantly underweight
    - Moderately underweight
    - Average weight
    - Moderately overweight
    - Significantly overweight

- What is the ethnicity of your biological mother?

- What is the ethnicity of your biological father?
